# Quantifying respiratory tract deposition of airborne graphene nanoplatelets: The impact of plate-like shape and folded structure

**DOI:** 10.1101/2020.08.28.20183608

**Authors:** Hanchao Gao, Tobias Hammer, Xiaole Zhang, Weidong He, Guangbiao Xu, Jing Wang

**Affiliations:** Key Laboratory of Textile Science & Technology, Ministry of Education, College of Textiles, Donghua University, No. 2999 North Renmin Road, Songjiang, Shanghai 201620, China; Institute of Environmental Engineering, ETH Zurich, 8093 Zurich, Switzerland; Advanced Analytical Technologies, Empa, Ueberlandstrasse 129, 8600 Dübendorf, Switzerland

**Keywords:** Graphene nanoplatelets, Aerodynamic diameter, Lung deposition, Multiple-path particle dosimetry model, Exposure assessment

## Abstract

The booming development of commercial products containing graphene nanoplatelets (GNPs) triggers growing concerns over their release into the air. Precise prediction of human respiratory system deposition of airborne GNPs, especially in alveolar region, is very important for inhalation exposure assessment. In this study, the pulmonary deposition of airborne GNPs was predicted by the multiple-path particle dosimetry (MPPD) model with consideration of GNPs plate-like shape and folded structure effect. Different equivalent diameters of GNPs were derived and utilized to describe different deposition mechanisms in the MPPD model. Both of small GNPs (geometric lateral size *d_g_* < 0.1 μm) and large GNPs (*d_g_* > 10 μm) had high deposition fractions in human respiratory system. The total deposition fractions for 0.1 μm and 30 μm GNPs were 41.6% and 75.6%, respectively. Most of the small GNPs deposited in the alveolar region, while the large GNPs deposited in the head airways. The aerodynamic diameter of GNPs was much smaller than the geometric lateral dimension due to the nanoscale thickness. For GNPs with geometric lateral size of 30 μm, the aerodynamic diameter was 2.98 μm. The small aerodynamic diameter of plate-like GNPs enabled deposition in the alveolar region, and folded GNPs had higher alveolar deposition than planar GNPs. Heavy breathing led to higher GNPs deposition fraction in head airways and lower deposition fractions in the alveolar region than resting breathing. Our results reveal that large GNPs can have small enough aerodynamic diameters to be respirable and deposit beyond the ciliated airways. The plate-like morphology and folded structure of GNPs resulted in higher alveolar deposition compared to spherical particles.

## 1. Introduction

Graphene nanoplatelets (GNPs) are two-dimensional nanoparticles made from graphite with typical lateral dimension of 0.5 – 20 μm and 0.34 – 100 nm in thickness. Two-dimensional GNPs exhibit superior mechanical strength [1], excellent electrical conductivity [2] and higher thermal conductivity [3] compared to zero-dimensional fullerenes and one-dimensional nanotubes. GNPs have been widely used in electronics [4], sensors [5], composite materials [6], energy storage [7] and biotechnology [8], and the global market of graphene is predicted to reach $311.2 million by 2022 [9]. Due to the fact that some GNPs are inevitably released into the environment during product manufacturing, use and disposal processes, the concern about their potential impact on environment and human health has been growing [10-15]. Released GNPs in the airborne form are especially troubling due to their high mobility and the possibility of entering human body. Inhalation uptake is the most critical exposure route compared to other pathways, such as dermal adsorption and ingestion. The National Institute for Occupational Safety and Health (NIOSH) published a research about engineering control for nanoscale graphene platelets during manufacturing and handling processes in 2011 [16]. According to this report, airborne graphene platelets with number concentration higher than 2 × 10^6^ cm^−3^ were found in the production areas. Compared with spherical particles with the same volume, GNPs have larger lateral dimensions owing to the plate-like shape. Attempted uptake of GNPs by macrophages could lead to frustrated phagocytosis and result in inflammation [17, 18]. Therefore, precise prediction of respiratory deposition of airborne GNPs is very important for human exposure risk assessment.

The shape of a particle is an important factor which influences the particle aerodynamic properties. The GNPs exhibit unique aerodynamics owing to the platelet morphology, nevertheless, studies about the plate-like shape impact on GNPs lung deposition are scarce. There are some researches on inhalation exposure of fiber-like shaped particles, such as carbon nanotubes (CNTs) [19, 20] and asbestos [21, 22]. As fiber-like particles tend to align with the airflow when inhaled into the respiratory tract, the length and aspect ratio of the particle are very important factors. The significance of particle length in asbestos toxicity was first studied by Vorwald et al. [23], subsequently the inflammatory response of fiber-like particles was confirmed by many studies [24, 25]. However, up till now, researches on the inhalation exposure of two-dimensional GNPs are far from adequate and how the plate-like structure affects the respiratory deposition is not clear.

Because of the large lateral dimension to thickness ratio and out-of-plane flexibility, GNPs are easily warped in the out-of-plane direction. This unique property and van der Waals attraction between graphene sheets facilitate the self-folded configuration [26]. The study about folding could be traced back to the 1990s, where folding was observed due to the friction between scanning probe microscopy tips and the surface of pyrolytic graphite. The mechanical stimulation overcomes the potential barriers for deformation and triggers self-folding, and the van der Waals attraction determines the stability of the self-folded pattern [27]. Recently, self-folding behavior of GNPs has been studied by more and more researchers. Cranford et al. [28] utilized an atomistic model to simulate the self-folding of graphene sheets and derived the critical self-folding length. Meng et al. [29] proposed a theoretical model based on finite deformation beam theory to predict the self-folding of graphene, and the theoretical model showed good agreement with molecular dynamics simulation. Folding converts GNPs into more complex shapes and affects GNPs transport dynamics during respiratory deposition. Therefore, it is important to take folding into consideration for more accurate lung deposition prediction of airborne GNPs.

The potential health impact of GNPs inhalation exposure has attracted substantial interest. The hazard of inhaled particles depends on their deposition site in the respiratory tract. Compared to particles in the bronchus which can be eliminated from sputum, particles in the alveolar are engulfed by the macrophages and cleared over several months, which makes them more harmful to human health [30]. Animal exposure experiments including intratracheal instillation [31], pharyngeal aspiration [18], and inhalation exposure [32] have been conducted to evaluate respiratory tract deposition of graphene related materials in the literature. Intratracheal instillation in mice resulted in pulmonary edema and dose-dependent acute lung inflammation, and 47% GNPs still in the lung after 4 weeks [33]. However, animal exposure experiments are limited to the evaluation of particle deposition in large bronchial airway, with less accuracy for small airway such as alveolar region. Lung deposition model provides an effective way to give more information about small airway deposition. The multiple-path particle dosimetry (MPPD) model and International Commission on Radiological Protection (ICRP) model were developed for simulating particle pulmonary deposition. Anjilvel et al. [34] firstly introduced the multiple-path model to estimate particle deposition in the rat lung. Subsequently, this model was improved and proved to be a reliable model to evaluate particle deposition in animals and humans [35, 36]. The MPPD model could simulate the deposition of non-spherical particles using particle equivalent diameters instead of geometric diameter to consider particle shape effect. As far as we know, there have been no studies of comprehensive respiratory deposition assessment of airborne GNPs using the MPPD model.

In the present work, the airborne GNPs deposition in the head airways, tracheobronchial and alveolar regions were simulated by the MPPD model. The GNPs aerodynamic diameter was derived based on the aerodynamics of oblate spheroids with random orientation. Various equivalent diameters (aerodynamic diameter, sedimentation diameter, mobility diameter) of GNPs were applied in the MPPD model to consider the plate-like shape impact on respiratory tract deposition. The folding effect was investigated by comparing the deposition fractions of planar GNPs and folded GNPs. In addition, the GNPs respiratory deposition was systematically discussed based on different breathing scenarios and respiratory parameters for comprehensive exposure assessment of airborne GNPs. This study not only revealed the aerodynamic characteristic, but also predicted the pulmonary deposition of GNPs and highlighted the impact of plate-like morphology and folded structure on higher alveolar deposition than spherical particles.

## 2. Results

### 2.1 Characterization of plate-like shape of folded structure of airborne GNPs

The pristine GNPs showed platelet shape with average lateral size of 5 μm (Fig. 1a). Airborne GNPs were produced by atomizing 0.02 wt% GNPs suspension and collected on Nuclepore membranes for geometric size measurement. The airborne GNPs remained the plate-like morphology (Fig. 1b), and the geometric lateral size of airborne GNPs was 0.5 − 3.5 μm measured from SEM pictures (Fig. 1c). The aerodynamic diameter of airborne GNPs was 0.54 − 1.4 μm detected by the Aerodynamic Particle Sizer (APS) (Fig. 1d). The minimum measurement threshold of APS was 0.54 μm, therefore the aerodynamic diameter measured by APS began from 0.54 μm. For both the geometric lateral diameter and aerodynamic diameter, the frequency distributions exhibited skewed shapes with long tails at large particle sizes, and the frequency distributions were well fitted to the lognormal distribution. The thickness of airborne GNPs was also quantified from high-resolution SEM pictures. As our samples were GNPs containing stacks of graphene sheets, the thicknesses of all GNPs were not the same but represented by a distribution. The measured thickness was well fitted by the normal distribution and thickness expectation was 66 nm (Fig. S1).

**Fig. 1.**
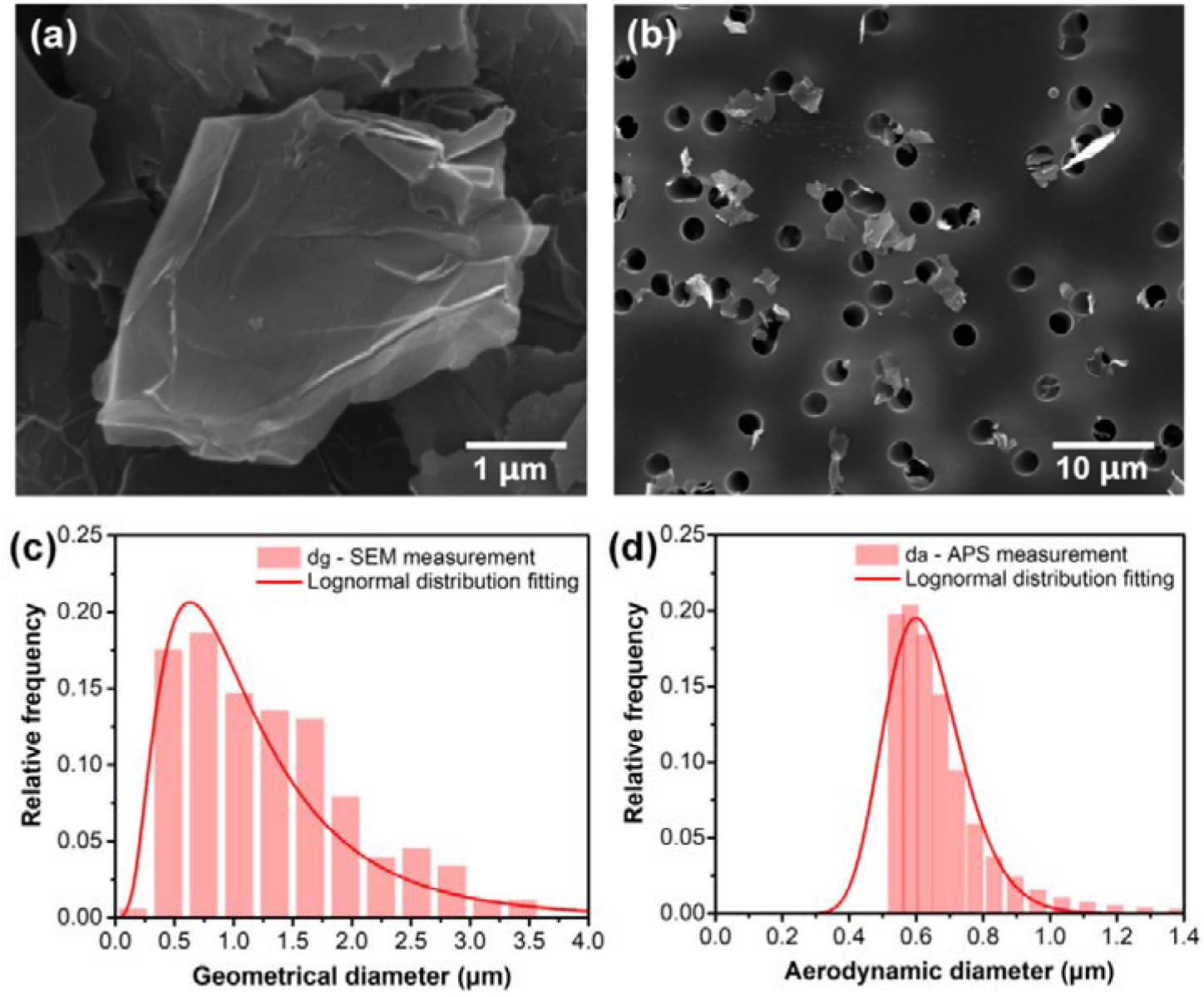
(a) SEM image of pristine GNPs, (b) SEM image of airborne GNPs collected on Nuclepore membrane, (c) geometric diameter frequency distribution of airborne GNPs from SEM measurement and fitted by lognormal distribution, (b) aerodynamic diameter frequency distribution of airborne GNPs from APS measurement and fitted by lognormal distribution.

Not only planar GNPs, folded GNPs were also observed from SEM and AFM images. Fig. 2a shows an airborne GNP with planar structure captured on the pore opening of the Nuclepore filter, and the planar GNP had plate-like shape with relatively flat surface (Fig. 2b-c). Fig. 2d reveals the characteristic of a folded GNP and an obvious bending e*d_g_*e is shown in the AFM height profile (Fig. 2e-f). The folded structure was owing to the out-of-plane flexibility and large lateral dimension to thickness ratio of GNPs. Folding was triggered by mechanical stimulus and stabilized via van der Waals forces [26, 27]. The folding frequency based on GNPs lateral size was also estimated. The folded GNPs were picked out manually in the SEM images (Fig. S2), and the numbers of folded GNPs in different size ranges were counted and corresponding folding frequencies were calculated (Table S1). There were also some factors influencing the accuracy of SEM images analysis, such as particles out of focus, overlapping particles and operator discretion. The number of such GNPs were also counted and added as the uncertainty of the folding frequency. The folding frequency increased with the geometric diameter from 14.3% for 0 – 1 μm GNPs to 27.4% for 2 – 3 μm GNPs. Larger lateral size not only affected the torque and made GNPs more deformable, but also resulted in larger adhesion energy to balance the bending energy and made folding states energetically stable [37]. During inhalation the incoming air negotiated a series of direction changes and respiratory deposition occurred in a complex system with inconstant flow rate. The flow velocity decreased dramatically from the lobar bronchi to the bronchioles due to the tremendously increased total cross-sectional area of numerous small airways [38]. The shear stress in the non-uniform and possibly turbulent airflow during inhalation could induce GNPs folding. Folding made GNPs into more intricate shape and influenced particle transport dynamics during respiratory deposition.

**Fig. 2.**
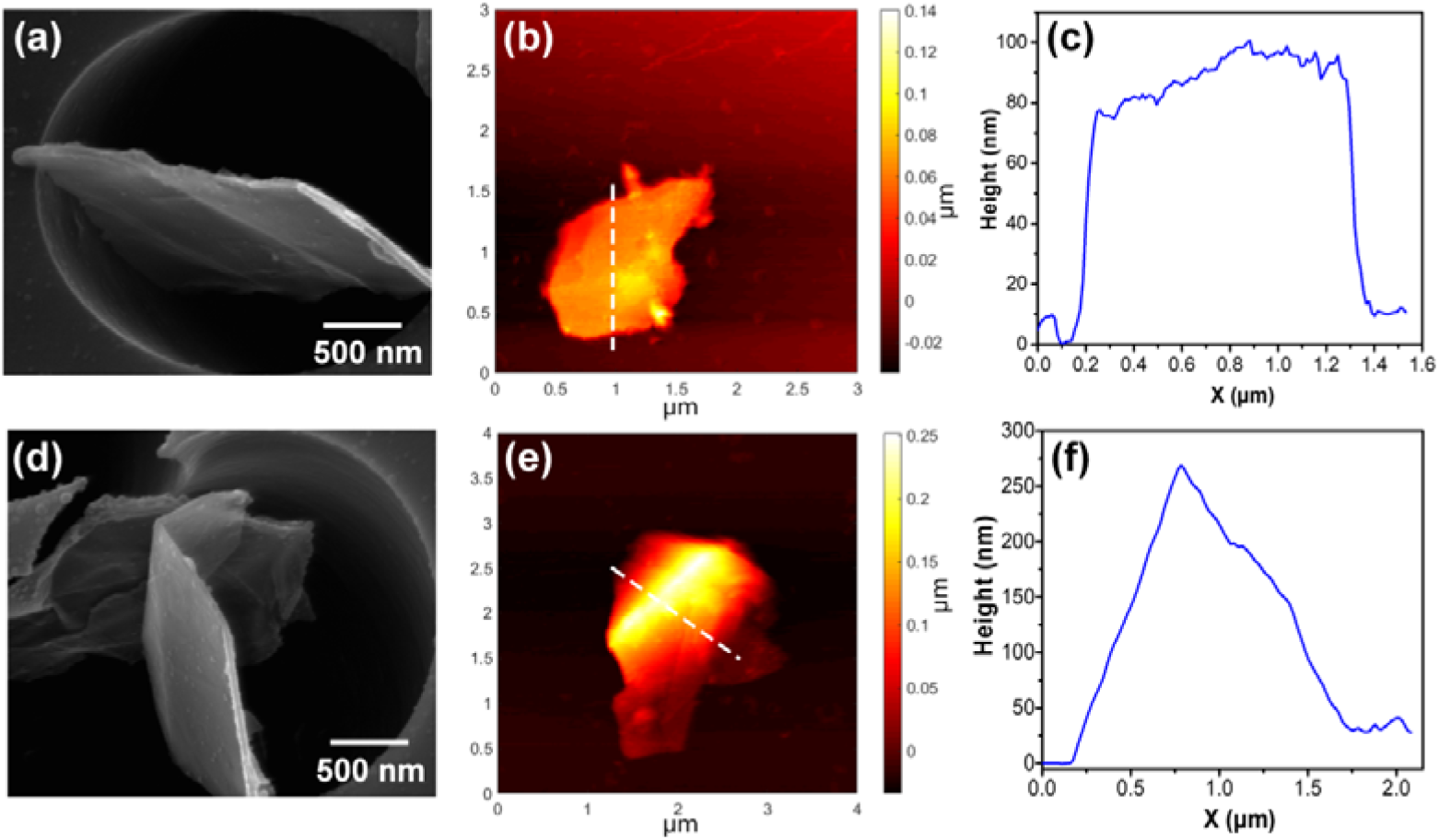
(a) SEM image of a planar GNP, (b) AFM image of a planar GNP, (c) height profile of the GNP corresponding to the dashed line in panel (b), (d) SEM image of a folded GNP, (e) AFM image of a folded GNP, (f) height profile of the GNP corresponding to the dashed line in panel (e).

### 2.2 The aerodynamics and equivalent diameters of GNPs

The transport behavior of plate-like particles could not be described by a single dimension such as the geometric diameter, as for spherical particles. The equivalent diameters (aerodynamic diameter *d_a_*, sedimentation diameter *d_s_*, mobility diameter *d_m_*) were important and used in the lung deposition model to describe different deposition mechanisms. The aerodynamic diameter *d_a_* is very significant as it determines the respirability of a particle and the site of deposition. Firstly, the calculated *d_a_* of GNPs was compared to the *d_a_* measured by APS to evaluate the derivation consistency. The measured *d_a_* was between the calculated *d_a_* of planar GNPs and folded GNPs, manifesting the analytical model calculation was reasonable (Fig. S3). The comparison among different equivalent diameters of GNPs is shown in Fig. 3a. For a GNP with particle volume of 1 μm^3^, the *d_g_*, *d_m_*, *d_a_* and *d_s_* were 4.4 μm, 3.32 μm, 1.14 μm and 0.77 μm. The mobility diameter *d_m_* was slightly smaller than the geometric lateral size *d_g_*, while the aerodynamic diameter *d_a_* and sedimentation diameter *d_s_* were much smaller than *d_g_*. The disparity among different equivalent diameters enlarged with increasing GNPs size. Numerous studies [39, 40] revealed that fiber-like particles possessed small aerodynamic diameter compared to geometric length. The aerodynamic diameter of glass fibers (fiber length: 30 μm, fiber diameter: 1 μm) was 3.47 μm [41], which was much smaller than the fiber length. Our results showed that fiber-like particles were not the only extended particles with small *d_a_*, the plate-like shaped GNPs also had small aerodynamic diameter in comparison with geometric lateral size.

**Fig. 3.**
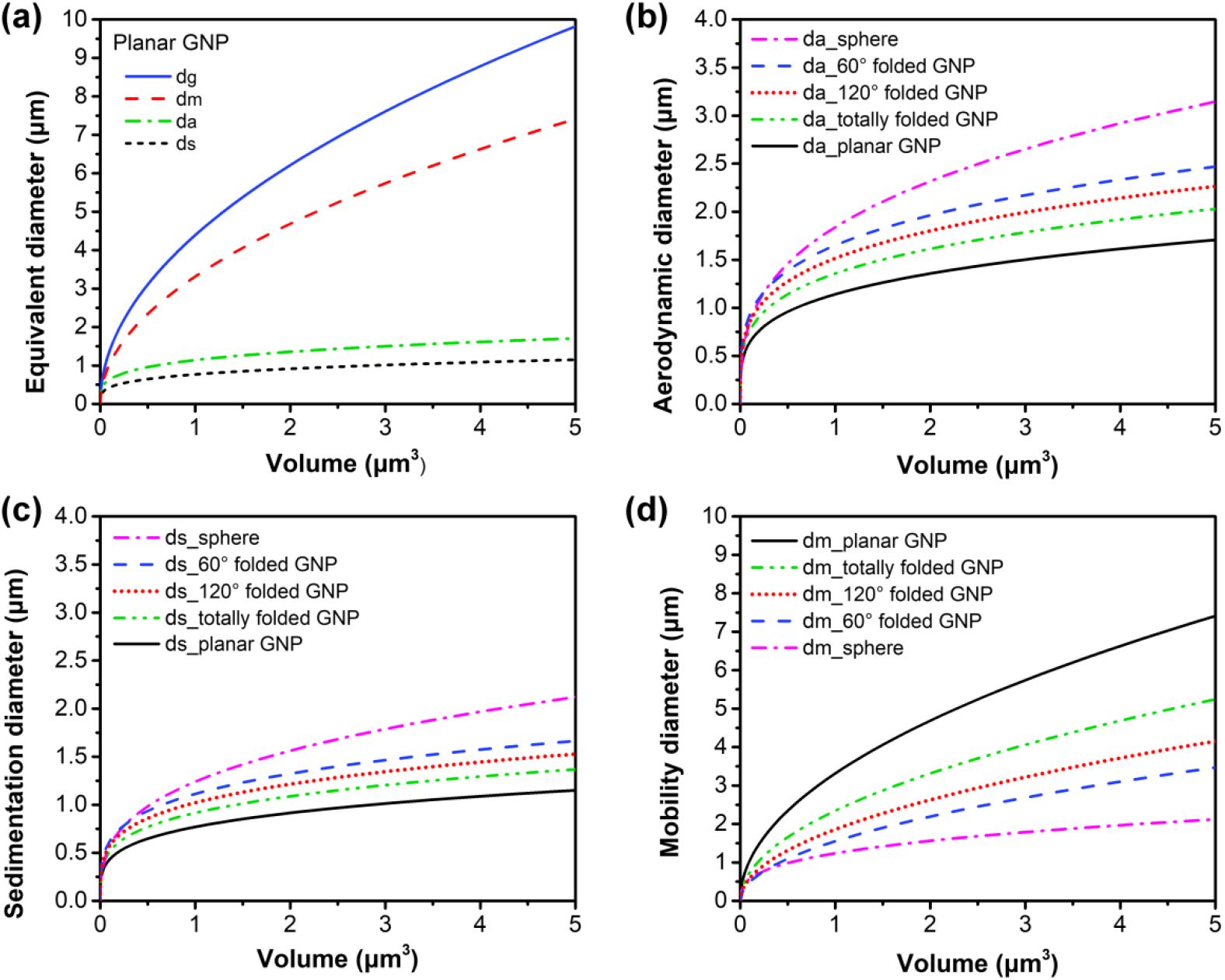
Comparison of (a) various equivalent diameters of planar GNPs, Comparison of (b) aerodynamic diameters *d_a_*, (c) sedimentation diameters *d_s_*, (d) mobility diameters *d_m_*, among planar GNPs, folded GNPs and spherical particles.

For GNPs with folding angles of 60º and 120º, firstly the drag forces were derived by numerical simulation instead of analytical expression, and then the equivalent diameters were calculated. The drag forces were simulated by integrating the pressure and viscous force on the GNP surface. The pressure and velocity distributions around GNPs with folding angles of 60º and 120º are shown in Fig. 4. The pressure distribution indicated that the windward side of the particle had higher pressure than the leeward side, which induced the drag force due to pressure. Compared to the air velocity near the boundary, the air velocity close to the particle was lower due to the viscosity, and the viscous force was calculated based on the air velocity gradients. The drag forces of GNPs with perpendicular and parallel orientations were simulated respectively, and the detailed numerical simulation results are shown in Table S2. The drag force of GNPs with perpendicular orientation was higher than that for parallel orientation. The drag forces of GNPs with folding angles of 60º and 120º were within the range of the drag forces of planar GNPs and totally folded GNPs, manifesting the simulation results were reasonable (Fig. S4).

**Fig. 4.**
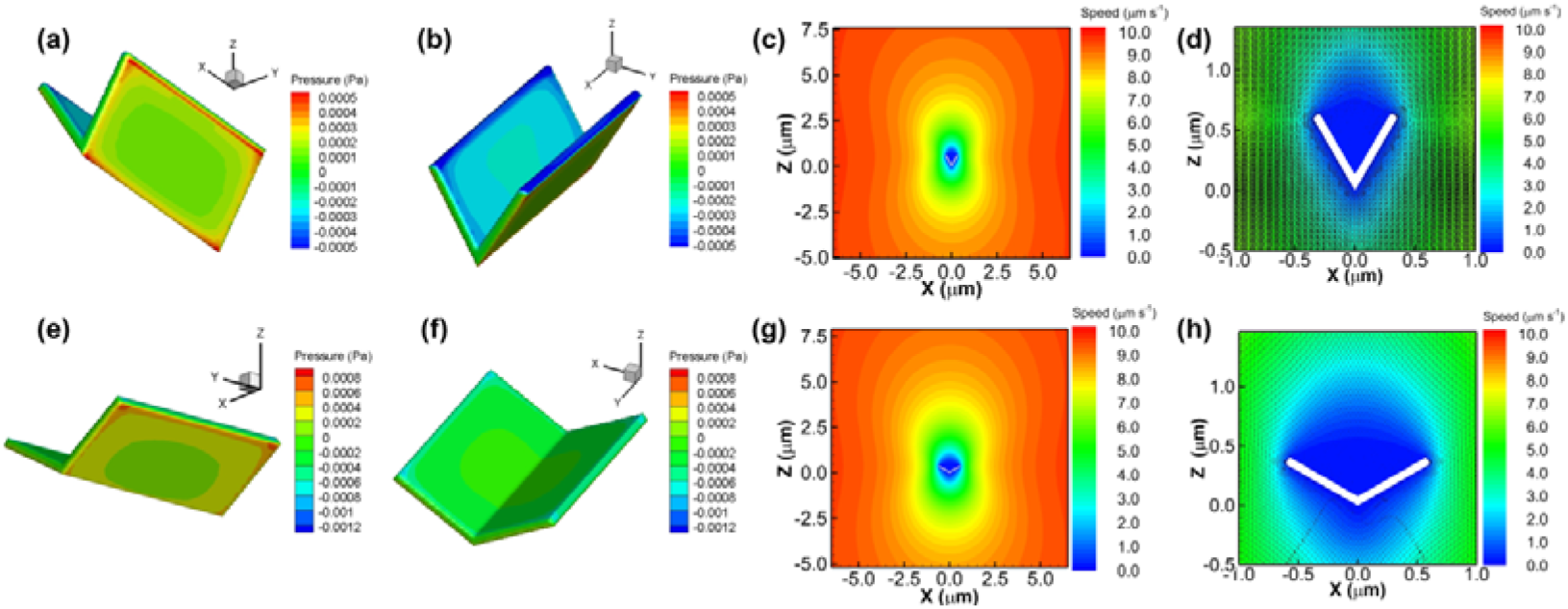
Pressure and velocity distributions around GNPs with folding angles of 60º and 120º. (a-b) pressure distribution and (c-d) velocity distribution (airflow velocity: 10^−5^ m s^−1^) around GNPs with folding angle of 60º and perpendicular orientation, (e-f) pressure distribution and (g-h) velocity distribution (airflow velocity: 10^−5^ m s^−1^) around GNPs with folding angle of 120º and perpendicular orientation.

Furthermore, the equivalent diameters comparison between GNPs and spherical particles was analyzed. The aerodynamic diameter *d_a_* and sedimentation diameter *d_s_* of GNPs were much smaller than those of spherical particles with the same volume (Fig. 3b-c), while GNPs had larger mobility diameter *d_m_* than spherical particles (Fig. 3d). The equivalent diameters disparity between folded GNPs and planar GNPs was highlighted to reveal the folding impact. A totally folded GNP was assumed having doubled thickness and half surface area of a planar GNP. Totally folded GNPs had larger aerodynamic diameter, larger sedimentation diameter, and smaller mobility diameter than planar GNPs with the same volume. The equivalent diameters of GNPs with folding angle of 60º and 120º were between the diameters of totally folded GNPs and spheres. Compared to 120º folded GNPs, the equivalent diameters of 60º folded GNPs were closer to the equivalent diameters of spheres. Different equivalent diameters of GNPs were used to depict different deposition mechanisms in the lung deposition model. As impaction was influenced by particle aerodynamics, the aerodynamic diameter *d_a_* was applied in the MPPD model for impaction deposition calculation. Instead of aerodynamic diameter, sedimentation deposition was determined by the sedimentation diameter *d_s_*, and the mobility diameter *d_m_* was more suitable for diffusion deposition. The plate-like shape and folded structure affected GNPs aerodynamic characteristic, further affecting GNPs pulmonary deposition. More analysis about the impact of GNPs plate-like shape and folded structure on respiratory tract deposition is discussed in the following section.

### 2.3 Respiratory deposition assessment of GNPs based on MPPD model

#### 2.3.1 Total and regional respiratory deposition of GNPs

The GNPs deposition in human respiratory system was simulated by the MPPD model. The total and regional deposition fractions *vs*. GNPs geometric diameter from 0.1 to 30 μm is demonstrated in Fig. 5. Three deposition mechanisms including impaction, sedimentation and diffusion were considered during respiratory deposition. As the airways structures were different in the head airway, tracheobronchial (TB) and alveolar regions, the dominant deposition mechanisms were also different in different regions [38]. The airway structure in head airways was complicated and airway diameter was wider and airflow rate was faster than other regions, therefore inertial impaction was the dominant mechanism for head airways. When the airflow containing GNPs passed through the airways, the tortuous airway structure affected the airflow trajectory and induced the change of airflow direction. Nevertheless, the GNPs were difficult to follow the streamline direction due to large momentum and detached from the streamline under inertia effect, and hit the bends and tortuous part of the airways. The momentum enhanced with GNPs diameter, resulting in the increased inertial impaction deposition with GNPs diameter. The head airway depositions of 1 μm and 30 μm GNPs were 7.4% and 57.0% with inhalation flowrate of 7.5 L min^−1^. Sedimentation and diffusion mechanisms were more significant in the TB and alveolar regions, where flow rate were low, residence times were long and airway dimensions were small. Diffusion deposition was effective for small particles. The deposition fractions of 0.1 μm GNPs in the TB region and alveolar region were 13.2% and 21.0%. Sedimentation deposition was more efficient for big particles and enhanced with particle size increase. However, as most of large GNPs were already deposited in the pharynx and larynx airways, the deposition fraction of large GNPs in the TB and alveolar regions did not increase obviously. The deposition fractions of 30 μm GNPs in the TB region and alveolar region were 5.2% and 13.4%. These results revealed that GNPs with large lateral size still could transit down to the alveolar region due to the small aerodynamic diameter. The alveolus did not have protective mucus layer and the only clearance mechanism of GNPs deposited in the alveolus was engulfed by alveolar macrophages which needed several months.

**Fig. 5.**
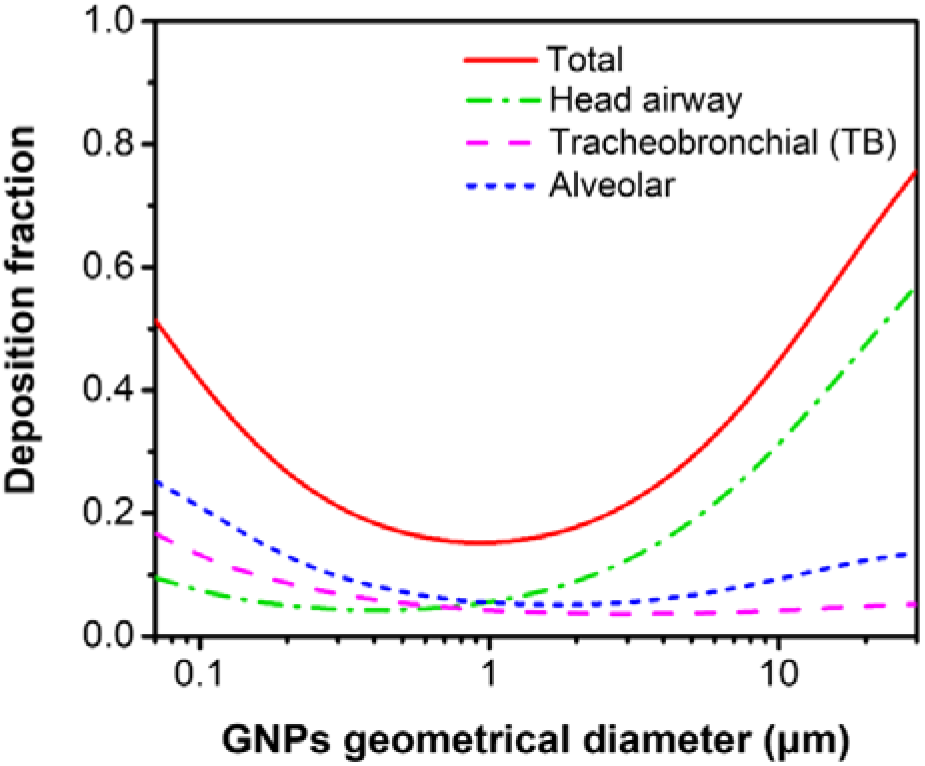
Total and regional deposition fractions of planar GNPs in human respiratory system with inhalation flowrate of 7.5 L min^−1^.

The total deposition fraction was the sum of deposition fractions in the head airway, TB and alveolar regions. As the diffusion mechanism was efficient for GNPs smaller than 0.1 μm, resulting in high alveolar deposition, thus the total deposition was high for these small GNPs. The total deposition fraction of 0.1 μm GNPs was 41.6%. With the GNPs size increase, the diffusion deposition was weakened, while the inertial impaction and sedimentation were enhanced, therefore the total deposition for large GNPs was high. The total deposition fraction of 30 μm GNPs was 75.6%. The GNPs diameter that gave the minimum total deposition, about 1 μm in Fig. 5, was the diameter that was too large for diffusion to be effective and too small for impaction and sedimentation to be effective. The total deposition fraction of 1 μm GNPs was only 15.3%.

#### 2.3.2 Deposition comparison between GNPs and spherical particles

Fig. 6 shows the human respiratory deposition comparison between GNPs and spherical particles. For small particles less than 0.3 μm, diffusion was the predominant deposition mechanism and particle shape effect was insignificant, thus the total deposition fraction was comparable for GNPs and spheres less than 0.3 μm. For GNPs and spheres between 0.3 and 16 μm, the total deposition fraction of GNPs was lower than spheres, nevertheless, for particles between 16 and 30 μm the GNPs deposition was higher than spheres. Spherical particles larger than 10 μm did not reach the TB and alveolar regions, however, due to the small aerodynamic diameter, 10 μm GNPs still could reach the alveolar region and showed 9.5% alveolar deposition.

**Fig. 6.**
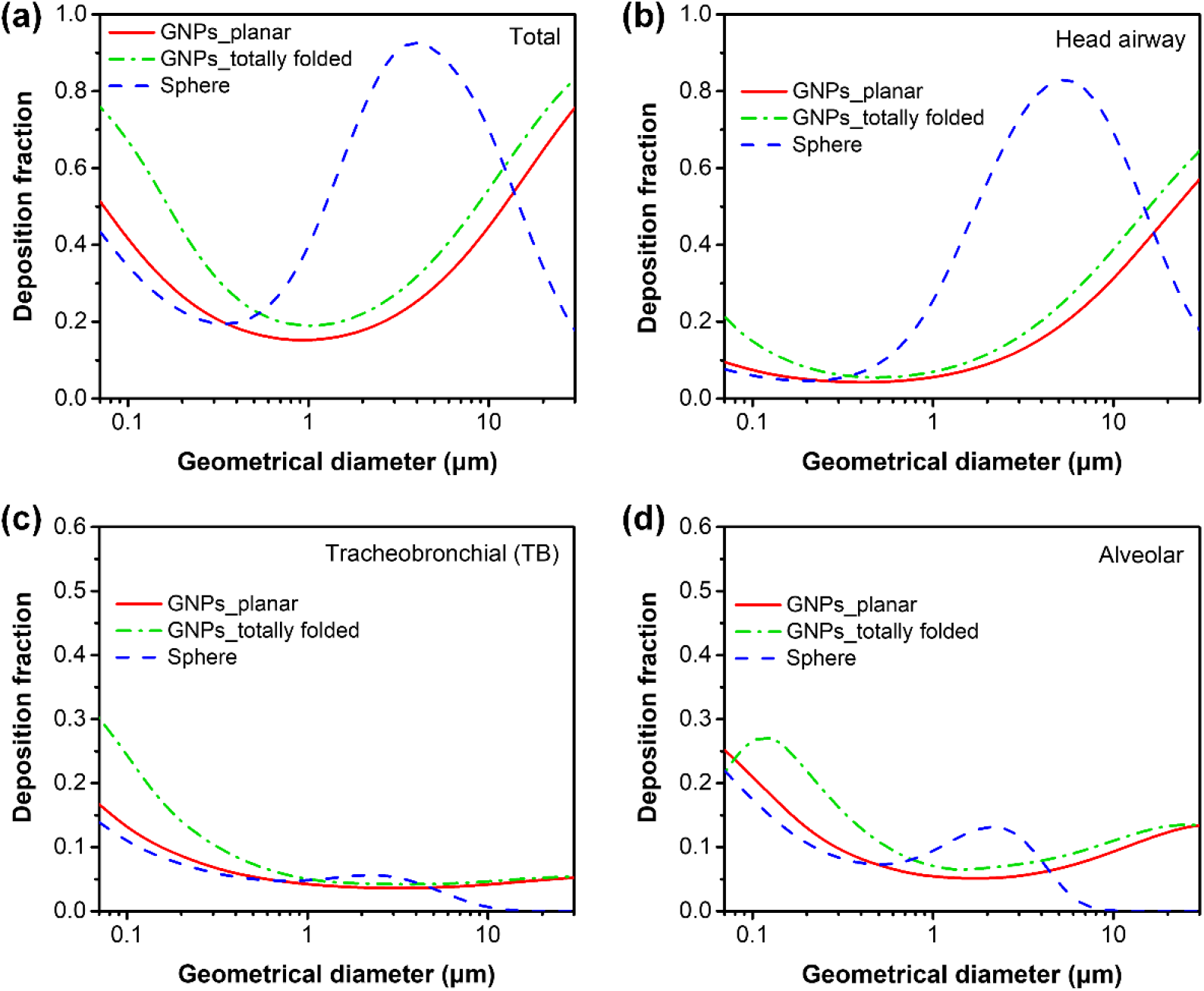
Comparison of the respiratory deposition between GNPs and spherical particles in human (inhalation flowrate: 7.5 L min^−1^), (a) total deposition, (b) head airway deposition, (c) TB deposition, (d) alveolar deposition.

Furthermore, the deposition fractions of planar GNPs and folded GNPs were compared to investigate the folding impact. Compared to planar GNPs with same lateral dimension, folded GNPs had larger aerodynamic diameter, larger sedimentation diameter, and smaller mobility diameter, which resulted in higher impaction, sedimentation and diffusion deposition. As impaction was the dominant mechanism in head airway, folded GNPs had higher head airway deposition due to the larger aerodynamic diameter than planar GNPs. As diffusion and sedimentation were efficient in the TB and alveolar regions, folding also resulted in higher TB and alveolar deposition owing to the larger sedimentation diameter and smaller mobility diameter. Therefore, for GNPs between 0.1 to 30 μm, folded GNPs had higher total deposition than planar GNPs.

#### 2.3.3 Effects of breathing scenarios and respiratory parameters on GNPs deposition

The location of inhaled particles depended not only on particle characteristics, but also on the individual’s breathing scenarios. Two different breathing scenarios including resting breathing (tidal volume: 625 ml, breath frequency: 12 per minute, flowrate: 7.5 L min^−1^) and heavy breathing (tidal volume: 1500 ml, breath frequency: 25 per minute, flowrate: 37.5 L min^−1^) of human were simulated by the MPPD model. The influences of flowrate on different mechanisms were different. The impaction deposition increased at higher flowrate. As impaction was the dominant mechanism for large particles deposited in head airways, the GNPs deposition in head airway was higher at heavy breathing (Fig. 7b). Similar trend was also reported by Sturm [42] who reported an obvious growth of GNPs head airway deposition with the inhalation flowrate increasing from 250 to 1000 cm^3^ s^−1^. On the other hand, high flowrate also meant short residence time and resulted in reduced diffusion and sedimentation, thus the GNPs deposition fractions in TB and alveolar regions decreased at heavy breathing (Fig. 7c-d). However, it should be noted that even though the deposition fraction was lower in the TB and alveolar regions at heavy breathing, as the flowrate of heavy breathing was five times that of resting breathing, the deposited amount at heavy breathing might still be higher with the same breathing time. Heavy breathing resulted in higher GNPs deposition in head airway, and lower deposition in TB and alveolar regions compared to resting breathing. The total deposition fraction was the sum of regional deposition fractions. For GNPs with *d_g_* < 0.2 μm the total deposition fraction under heavy breathing and resting breathing were similar. For GNPs with *d_g_* > 0.2 μm, as the increased head airway deposition overwhelmed the decreased TB and alveolar deposition, the total deposition fraction at heavy breathing was higher than resting breathing (Fig. 7a). The effect of breathing scenarios on the deposition fractions of GNPs and spheres were also compared. For spheres with geometric diameter between 0.1 to 30 μm, the deposition fractions in TB and alveolar regions at heavy breathing were lower than those at resting breathing. The head airway deposition for spheres with *d_g_* < 5 μm was higher at heavy breathing, whereas for spheres with *d_g_* > 5 μm the head airway deposition at heavy breathing was lower than resting breathing.

**Fig. 7.**
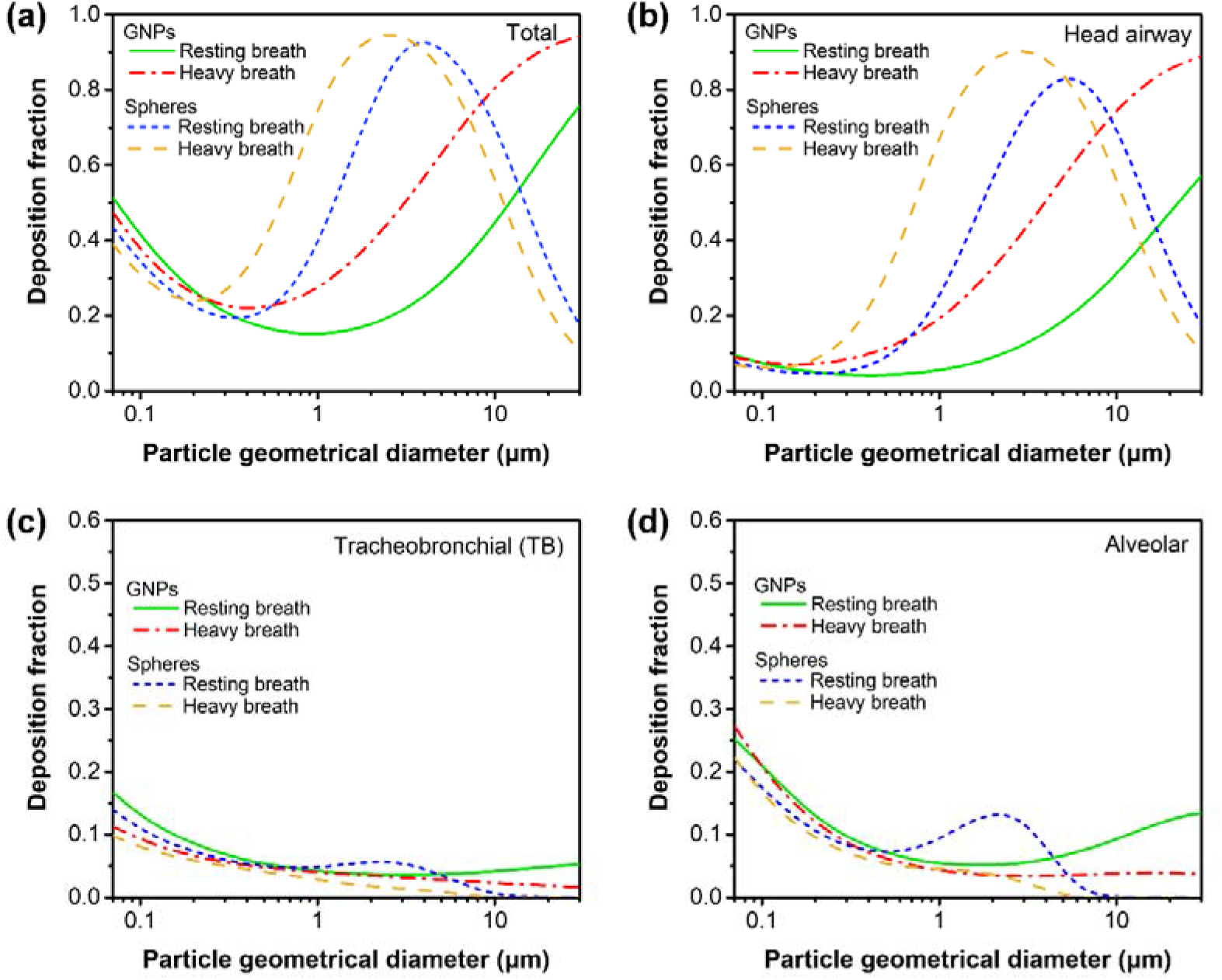
Comparison of respiratory deposition of planar GNPs and spherical particles between two different breathing scenarios: resting breathing (7.5 L min^−1^) and heavy breathing (37.5 L min^−1^), (a) total deposition, (b) head airway deposition, (c) TB deposition, (d) alveolar deposition.

Furthermore, the respiratory deposition of GNPs was compared between human and different animals (rat, pig and rabbit). Human had larger FRC and URT volumes, larger tidal volume and lower breath frequency than animals (Table S3). Due to the different lung structure and respiratory physiological parameters between human and animals, it was difficult to predict human respiratory deposition dosimetry from animal experiments. The MPPD model provided a biologically-based method to solve this problem and facilitate the determination of the suitable surrogate which had the most similar deposition fraction as human. Fig. 8a indicated rat had the most similar total deposition fraction as human, while the deposition fractions in pig and rabbit were lower, especially for GNPs larger than 1 μm. For head and TB regions, rat had the most similar deposition fractions as human (Fig. 8b-c). Nevertheless, for GNPs larger than 1 μm, all three kinds of animals had lower alveolar deposition than human (Fig. 8d). In addition, the respiratory deposition of spherical particles in human and animals were compared, and rat also showed the most similar deposition as human (Fig. S5). The plate-like structure of GNPs did not influence the selection of suitable surrogate. The graphene pulmonary deposition fractions comparison between rat and human calculated by MPPD model were also reported by Lee et al. [35]. The deposition fractions were quite similar between rat and human for the extrathoracic and tracheobronchial regions, nevertheless, the deposition fraction in the alveolar region of rat (0.0569) was lower than that of human (0.1043). The modeling results in our research matched well with the results in the literature, furthering confirming the validity of our results. In summary, for the deposition fraction consideration, rat would be the suitable surrogate animal for human exposure assessment, nevertheless, it should also be noticed that rat showed lower alveolar deposition than human for GNPs larger than 1 μm.

**Fig. 8.**
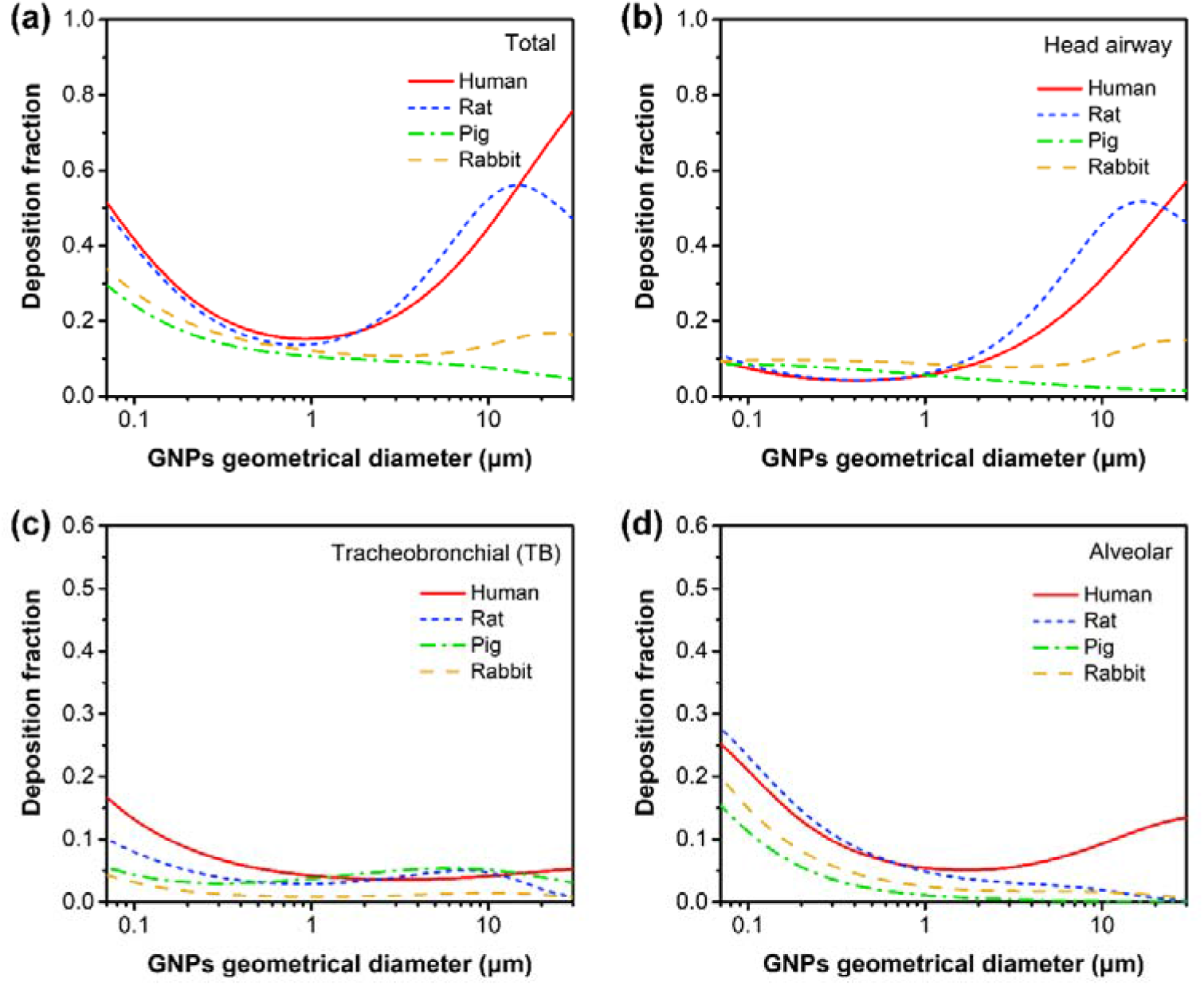
Comparison of planar GNPs respiratory deposition between human and different animals (rat, pig and rabbit), (a) total deposition, (b) head airway deposition, (c) TB deposition, (d) alveolar deposition.

## 3. Discussion

In this study, the aerodynamic diameter of GNPs was derived from the aerodynamics of oblate spheroids by considering the gravitational force and aerodynamic resistance perpendicular and parallel to particle motion, and the effect of random orientation was taken into consideration. For GNPs with 30 μm geometric lateral size, the aerodynamic diameter was 2.98 μm, which was within the respiratory particle size. The calculation results in our study matched well with the results in the literature. Sturm’s [42] derived the aerodynamic diameter of GNPs by analytical model and revealed for GNPs with 30 μm projected diameter, the aerodynamic diameter was 3.3 μm. Schinwald et al. [18] revealed the relationship between projected diameter and aerodynamic diameter of GNPs, and found for GNPs with 30 μm projected area diameter, the aerodynamic diameter was 3.26 μm. In addition, the dynamic shape factor was commonly used as a correction factor to explain the shape impact on particle motion. Sanchez et al. [17] derived the aerodynamic diameter of GNPs using the shape factor determined for oblate spheroids, and reported for GNPs with lateral size of 25 μm, the aerodynamic diameter was between 1.25 to 1.5 μm depending on the particle orientation. Our research and literature studies confirmed that the aerodynamic diameter of GNPs were smaller than the lateral dimension due to the nanoscale thickness. For GNPs with large lateral size, the aerodynamic diameter was still small and respirable. Apart from GNPs, there were plate-like minerals such as talc, mica and nanoclay. It was reported the aerodynamic diameter *d_a_* of talc was smaller than the projected area diameter *d_p_* [43]. The Stokes diameter of mica was smaller than the equivalent volume diameter [44]. Our research and above studies showed that fiber-like shaped particles were not the only case possessing small aerodynamic diameter, the plate-like shaped particles also had small aerodynamic diameter compared to the geometric lateral size.

Due to the small aerodynamic diameter, the GNPs with large geometric lateral size were still respirable and could deposit beyond the ciliated airways. The MPPD simulation showed the alveolar deposition fractions of 10 μm and 30 μm GNPs were 9.5% and 13.4% with the inhalation flowrate of 7.5 L min^−1^. Animal exposure experiments have been conducted to evaluate respiratory tract deposition of graphene related materials in the literature. A 5-day graphene inhalation toxicity study was executed in male rats by nose inhalation [45]. The rats were exposed to 3.86 mg m^−3^ graphene for 5 days, high-resolution dark-field imaging demonstrated the graphene deposited in the alveolar macrophages and accumulated in the rat lung. Rats Inhalation exposure experiments by Ma-Hock et al. [32] showed that after nose exposing to 10 mg m^−3^ graphite nanoplatelets (lateral size up to 30 μm, aerodynamic diameter about 2 μm) for 5 days, black irregularly shaped particles with lateral size about 5.6 μm were observed within the alveolar macrophages on day 4, these particles were still observed at day 95, and these black particles were regarded as graphite nanoplatelets. Mao et al. [33] analyzed the *in vivo* distribution of few layer graphene (lateral size: 60 − 590 nm) in mice after intratracheal instillation, most of graphene was remained in lung and 47% still maintained after 28 days. Graphene was found in the cytoplasm of alveolar macrophages, indicating graphene was phagocytized by alveolar macrophages after intratracheal instillation, and 0.15% and 1% graphene was redistributed to the spleen and liver via crossing the air-blood barrier. The above animal exposure experiment confirmed that the inhaled GNPs could deposit in the alveoli and phagocytized by alveolar macrophages. However, as the alveolus was located in the deep lung with micro-scale size, it was difficult to accurately obtain the alveolar deposition fraction from animal exposure experiments. The lung deposition model was commonly used to predict the deposition fraction, especially for the small airways. Apart from MPPD model, ICRP model was also utilized in previous research. The GNPs deposition calculated by ICRP model was reported by Sanchez et al. [17], indicating that the alveolar deposition fraction of 25 μm GNPs was about 10%. Sturm’s study [42] revealed that GNPs had higher alveolar deposition compared to spheres with equivalent volume, and the alveolar deposition fraction of 30 μm GNPs was 23%. This result was higher than our results which might be due to the different equations, as well as the different respiratory physiological parameters used in the model.

The lateral size played a significant role in biological interaction and determined the consequence of cellular uptake. The small aerodynamic diameter of GNPs enabled their deposition beyond the ciliated airways, as they still had large lateral size, they were too large to be completely phagocytosed by macrophages and could result in the inflammatory response. The inhalation toxicity of GNPs and carbon black was compared by pharyngeal aspiration in mice [18]. GNPs induced ganulomatous lesion in the alveolar region and pro-inflammatory cytokines in the bronchoalveolar lavage fluid, whereas carbon black did not induce inflammatory response. *In vitro* cell/particle interaction experiments showed that GNPs could not be fully engulfed by THP-1 cells and led to frustrated phagocytosis. It should also be noticed that the inhalation toxicity of carbon-based nanomaterials was quite complex. It was not only determined by dose or lateral size, but also depended on the combination of several physicochemical properties such as surface area, disperse state and durability. The inhalation toxicity of four different carbon-based nanomaterials (carbon black, MWCNTs, graphene, graphite nanoplatelets) was compared by rat inhalation exposure experiments, and toxicity was investigated by broncho-alveolar lavage fluid biochemical changes examinations [32]. MWCNTs and graphene generated lung toxicity, whereas no adverse toxicity was observed after exposure to graphite nanoplatelets or carbon black.

Compared to the previous research, this study had three novel contributions. Firstly, the lung deposition of folded GNPs was discussed for the first time. For GNPs between 0.1 to 30 µm, folded GNPs had higher alveolar deposition than planar GNPs. This result emphasized the need to consider the folding effect for more accurate dosimetry analysis of inhaled GNPs. Secondly, we not only analyzed the lung deposition of GNPs, but also compared these results with spherical particles to highlight the plate-like shape effect. The biggest deposition difference occurred in the TB and alveolar regions. Spherical particles larger than 10 μm could not reach the alveolar region, while GNPs with lateral size larger than 10 μm still could deposit in the alveolar region due to the small aerodynamic diameter. This characteristic deserved close attention as particles in the alveolar region were slowly cleared and more harmful for human health. Thirdly, the respiratory deposition of GNPs was comprehensively discussed based on different breathing scenarios and respiratory parameters. Heavy breathing brought out higher GNPs deposition fraction in head airway, and lower deposition fractions in TB and alveolar regions. The above information was helpful for comprehensive exposure assessment of airborne GNPs.

## 4. Conclusions

In summary, the plate-like morphology and folded structure affected the aerodynamic property and equivalent diameters of GNPs, further affecting GNPs transportation and deposition in the respiratory tract (Fig. 9). Both of small GNPs (*d_g_* < 0.1 μm) and large GNPs (*d_g_* > 10 μm) had high total deposition fractions in human respiratory tract calculated by the MPPD model. The total deposition fractions for 0.1 μm and 30 μm GNPs were 41.6% and 75.6%, respectively. Most of the small GNPs deposited in the alveolar region due to diffusion deposition, while the large GNPs had high head airway deposition owing to the dominant impaction deposition. The aerodynamic diameter of GNPs was much smaller than the lateral dimension due to the nanoscale thickness. For GNPs with geometric lateral size up to 30 μm, the aerodynamic diameter was 2.98 μm. The small aerodynamic diameter of GNPs enabled their deposition beyond the ciliated airways. The alveolar deposition fraction of 10 μm GNPs was 9.5%, and folded GNPs had higher alveolar deposition than planar GNPs. Heavy breathing brought out higher GNPs deposition fraction in head airway and lower deposition fractions in TB and alveolar regions than resting breathing.

**Fig. 9.**
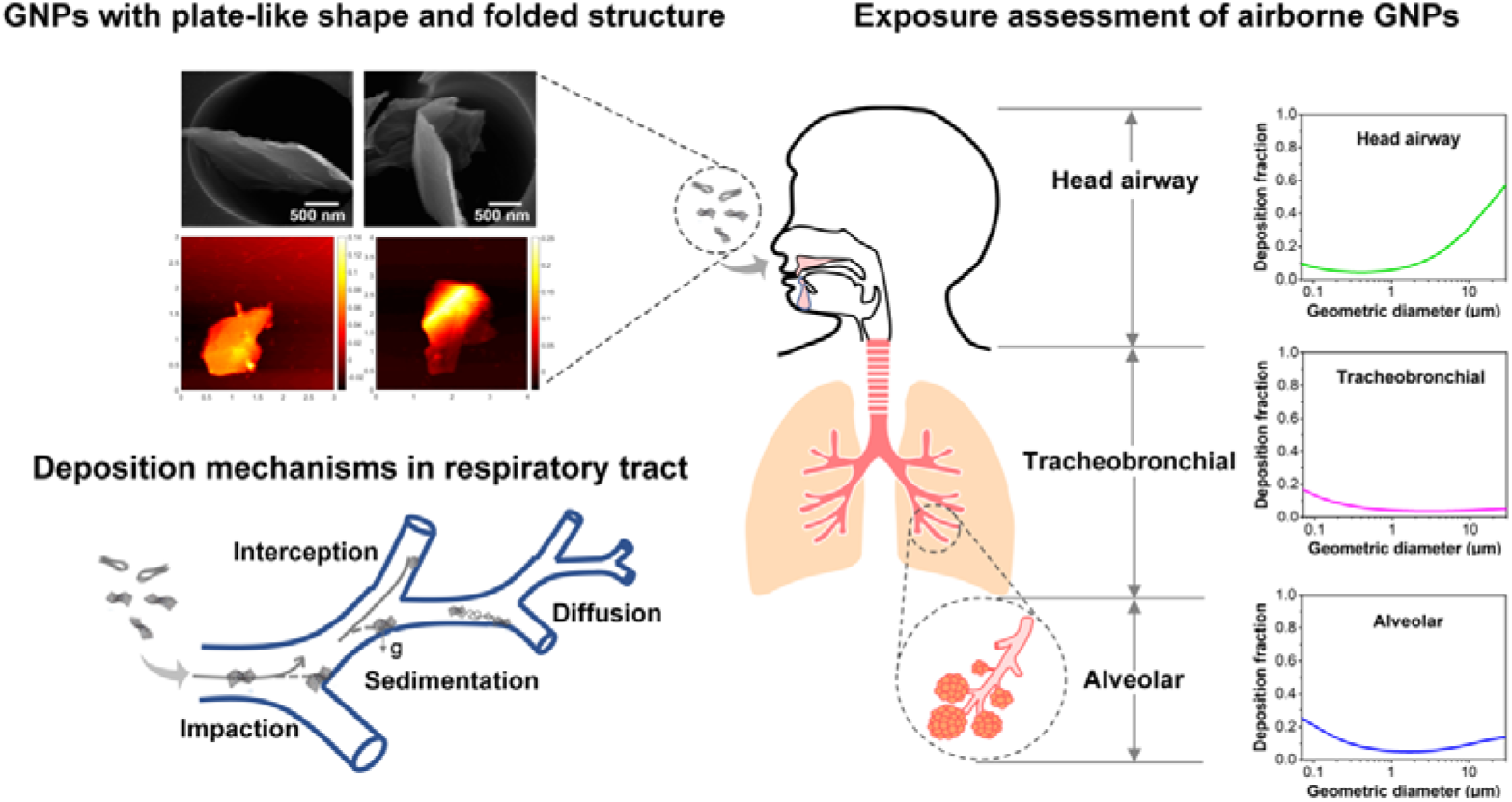
Summary illustration showing the plate-like and folded structures and respiratory deposition assessment of airborne GNPs.

## Methods

### Characterization of airborne GNPs

The graphene nanoplatelets (GNPs) were provided by XG Science, USA. The average geometric lateral dimension was 5 μm. The specific surface area was 120 − 150 m^2^ g^−1^ and the true density was 2.2 g cm^−3^. Airborne GNPs were produced via a Collison type atomizer by atomizing 0.02 wt% GNPs suspension. GNPs were dispersed in water and ultrasonicated for 20 min before atomizing to avoid agglomeration. Airborne GNPs were collected on Nuclepore membranes (WHA-111112, Whatman International, UK) for structure characterization and geometric size measurement. Nuclepore filters are thin polycarbonate films with uniform cylindrical holes perpendicular to the filter surface. The morphology, lateral size and thickness of airborne GNPs were measured via Scanning Electron Microscopy (SEM, Nova NanoSEM 230, Thermo Fisher Scientific, USA) and Atomic Force Microscopy (AFM, Solver Nano, NT-MDT Spectrum Instrument, Russia). The aerodynamic diameter of airborne GNPs was detected by the Aerodynamic Particle Sizer (APS, model 3321, TSI Inc., USA).

### Analytical expression for the equivalent diameters of planar GNPs and totally folded GNPs

#### Aerodynamic diameter *d_a_*

The aerodynamic diameter *d_a_* was significant for calculating particle deposition due to impaction mechanism. When a particle was released in air, it reached its settling velocity as the drag force offset the gravity. The drag force of plate-like GNPs was derived on basis of the drag force expressions for oblate spheroid [46], which has been commonly applied for plate-like particle aerodynamics calculation in previous studies [43, 47]. The aerodynamic drag force for GNPs depended on particle orientation. With the assumption that the particle thickness was infinitely thin, for a GNP perpendicular to the gas flow the drag force *F_D1_* was calculated as:

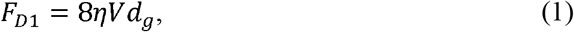

For a GNP parallel to the gas flow the drag force *F_D2_* was derived as:

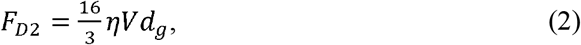

with *η*: gas viscosity, *V*: velocity of the GNP, *d_g_*: GNP geometric diameter.

The aerodynamic diameter of an irregular-shaped particle was defined as the diameter of a sphere with unit density and same terminal velocity as the irregular particle [38]. The aerodynamic diameter *d_a1_* of perpendicular orientated GNP and *d_a2_* of parallel orientated GNP were calculated as:

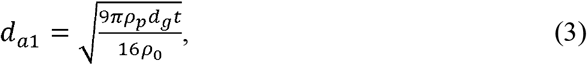

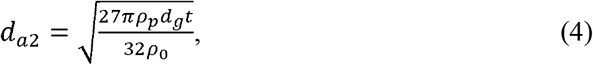

with *t:* thickness of GNPs*, ρ_0_*: unit density, *ρ_p_*: GNPs density (2.2 g cm^−3^).

The aerodynamic diameter *d_a_* of a random orientated GNP was derived as:

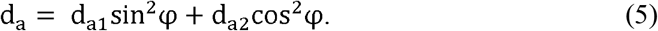

with *sin*^2^ *φ* = 2/3 and *cos*^2^ *φ* = 1/3 calculated by Fuchs [48] by balancing the resistance in all directions.

### Sedimentation diameter *d_s_*

The sedimentation diameter *d_s_* was defined as the diameter of a sphere with same density and equivalent terminal velocity as the irregular particle [38]. Compared with aerodynamic diameter which standardized for both shape and density, the sedimentation diameter *d_s_* only took the shape effect into account. Using the similar derivation method, the sedimentation diameter *d_s1_* of perpendicular orientated GNP and *d_s2_* of parallel orientated GNP were derived as:

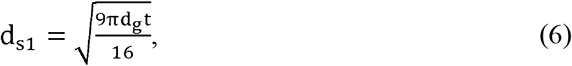

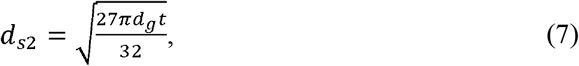

The sedimentation diameter *d_s_* of a GNP with random orientation was derived as:

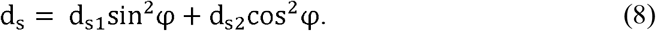

### Mobility diameter *d_m_*

The mobility diameter *d_m_* was significant for calculating particle deposition by diffusion mechanism. The mobility diameter *d_m_* was defined as the diameter of a sphere with equal terminal velocity as the irregular particle in the electric field [38]. A particle with *n* units charge was driven under the electrostatic force *F_E_* = *neE* in the electric field with field intensity *E*. The particle attained terminal velocity as the drag force offset electrostatic force. The mobility diameter *d_m1_* of perpendicular orientated GNP and *d_m2_* of parallel orientated GNP were derived as:

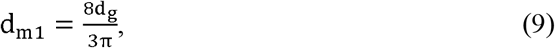

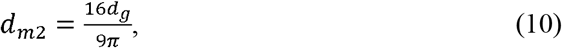

The mobility diameter *d_m_* of a random orientated GNP was derived as:

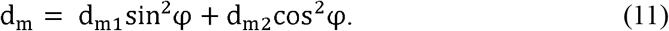

### Numerical simulation of drag forces of GNPs with folding angles of 60º and 120º

For GNPs with certain folding angles (e.g. 60**º** and 120**º**), it was difficult to derive the drag force from analytical expressions, nevertheless, the computational fluid dynamics (CFD) method provided a reliable way to obtain the drag force from numerical simulation. We adopted the SimpleFoam solver in the open source platform OpenFOAM Version 5.0. The geometry of the particle was generated using FreeCAD Version 0.18.1. The planar GNP was assumed to be a square cuboid, with a lateral size of 1.4 μm and a thickness of 66 nm. Folding was assumed to be along the symmetry axis of the GNP, and the center of the folding edge was located at (0, 0, 0). The size of the computation domain was 100 μm × 100 μm × 150 μm (x × y × z) (Fig. S6a). Grading mesh was utilized to generate finer grids around the particle. The grid was refined on the surface of the particle with about 0.02 μm in size near the particle (Fig. S6b). Ten additional layers were added near the particle surface with the minimum thickness of 0.001 μm to better calculate the velocity gradients and the viscous force (Fig. S6c). For the boundary conditions, the inlet flow came from the bottom boundary, which was defined as the inlet with a fixed velocity (e.g. (0, 0, 10^−5^) m s^−1^). The upper boundary was set as the fixed pressure condition. All the boundaries on the sides were defined as slip boundary. Since the side boundaries were far away from the particle (about 36 times the particle size), the influences of the boundary conditions should be insignificant. The drag forces of the particles were calculated with different face velocities at 10^−5^, 10^−4^ and 5 × 10^−4^ m s^−1^. The CFD results were first calibrated by the comparison with the Stokes’ law for the drag force on a sphere. The results agreed well with the Stokes’ law (Fig. S7). The relative errors were within 5%, which might come from approximation of the sphere geometry constructed by discrete grids. More details about the derivation of drag forces and the equivalent diameters of GNPs with folding angles of 60º and 120º are shown in the Supplementary Information.

### Calculation of GNPs lung deposition using the MPPD model

The pulmonary deposition of airborne GNPs was calculated by the multiple-path particle dosimetry (MPPD) model. The MPPD model is a simplified mathematical description of mechanisms and processes involved in deposition of inhaled particles as a function of key parameters such as particle characteristics, lung geometry and breathing scenario. This model predicts particle deposition in human and animal respiratory tracts with wide particle size range and series of breathing scenarios. The MPPD model could simulate the lung deposition of irregular shape particles using the particle equivalent diameters instead of geometric diameter. In the MPPD model, the human respiratory system is constructed using morphometric data compiled by Yeh and Schum [49] (Table S4). The respiratory system is divided into three regions. The first is the head airways region, which contains the nose, mouth, pharynx and larynx. The second is the tracheobronchial (TB) region, which includes the airways from trachea to terminal bronchioles. The third is the alveolar region. The inhaled particles deposit in different regions by diffusion, sedimentation and impaction. In the head region, the particle deposition was calculated using the method from Rudolf et al. [50] for impaction and Swift et al. [51] for diffusion. In the TB and alveolar regions, the particle deposition fraction was calculated based on Zhang et al. [52] for impaction, Ingham et al. [53] for diffusion and Wang et al. [54] for sedimentation. In this study, different GNPs equivalent diameters relevant to various deposition mechanisms (aerodynamic diameter for impaction deposition, mobility diameter for diffusion deposition, sedimentation diameter for sedimentation) were utilized in the MPPD model. The exposure condition was constant exposure with fixed breathing frequency and tidal volume (air respiration volume in a single breath), and the ratio between the inhalation time and total time was 0.5. The breathing scenario was nose exposure. In addition, the lung deposition of GNPs was also compared between human and different animals (rat, pig and rabbit) to determine the suitable surrogate animal that had the most similar deposition fraction as human. The detailed respiratory parameters for human and animals were summarized in previous study [55] and given in Table S3.

## Data Availability

All data and materials are included in this manuscript and the supplementary information file.

## Acknowledgements

This work is supported by Swiss National Science Foundation (grant number 310030_169207). The authors thank financial support from China Scholarship Council.

## Notes

### Competing Interest Statement

The authors have declared no competing interest.

### Author Declarations

No IRB necessary.

